# Detection of mTBI in pediatric populations using BrainCheck, a tablet-based cognitive testing software

**DOI:** 10.1101/2020.04.29.20085274

**Authors:** Siao Ye, Brian Ko, Huy Phi, David M. Eagleman, Benjamin Flores, Yael Katz, Bin Huang, Reza Hosseini Ghomi

## Abstract

**Aim:** Despite its high frequency of occurrence, mild traumatic brain injury (mTBI), or concussion, is difficult to recognize and diagnose, particularly in pediatric populations. Conventional methods to diagnose mTBI primarily rely on clinical questionnaires and sometimes include imaging such as computed tomography (CT) or pencil and paper neuropsychological testing. However, these methods are time consuming, require administration/interpretation from health professionals, and lack adequate test sensitivity and specificity. We explore the use of BrainCheck, a computerized neurocognitive test that is available on iPad, iPhone or computer desktop, for mTBI assessment. The BrainCheck battery consists of 6 gamified traditional neurocognitive tests that assess areas of cognition vulnerable to mTBI such as attention, processing speed, executing functioning, and coordination.

**Methods:** We administered BrainCheck to 10 participants diagnosed with mTBI at the emergency department (ED) of Children’s hospital within 96 hours of admittance to the ED, and 126 normal controls at a local high school. Statistical analysis included Chi-Square tests, Analysis of Variance (ANOVA), independent sample t-tests, and Hochberg tests to examine differences between mTBI, diagnoses by current gold standard clinical exam, and control groups on each assessment in the battery. Significant metrics from these assessments were used to build a logistic regression model that distinguishes mTBI from non-mTBI participants.

**Results:** BrainCheck was able to detect significant differences in coordination, Flanker, Stroop, Digital Symbol, Immediate/Delayed recall and recognition assessments between normal people and mTBI patients. Receiver operator score (ROC) analysis of our logistic regression model found a sensitivity of 84% and specificity of 80%.

**Conclusions:** BrainCheck has potential in distinguishing mTBI from non-mTBI participants, by providing a shorter, gamified test battery to assess cognitive function after brain injury, while also providing a method for tracking recovery with the opportunity to do so remotely from a patient’s home.

## Introduction

Mild traumatic brain injury (mTBI) is an increasing public health concern not only due to its growing frequency of occurrence but lack of guidelines and biomarkers that make diagnosis challenging [1]. Recent evidence also indicates an association to the development of neurodegenerative diseases [2]. This is particularly concerning in the pediatric population whose ongoing cognitive maturation makes them more vulnerable to head injury than the adult population [3]. The diagnostic utility of neuroimaging is limited and current recommendations do not recommend routine imaging for mTBI unless otherwise clinically indicated [1]. Furthermore, literature evaluating sports-related concussions (SRCS) has indicated that young athletes with mTBI may not recognize alarming symptoms due to a lack of awareness, understanding, or as a result of their cognitive impairment [4]. Thus, self-reported post-concussion symptoms are not reliable in this population. Additionally, while there are several concussion grading systems, there is little agreement between the systems on how they define and assess mTBI, and a collectively agreed gold-standard grading system does not exist [5]. Self-reporting concussion symptoms is often at odds with social pressures on a younger person to perform both educationally and athletically and places a strong bias making these measures more inaccurate [6].

Although neuropsychological tests were developed to more accurately measure cognitive deficits in mTBI, these tests have their limitations. Typically they require a neuropsychologist or psychometrist to administer, score, and interpret the results of the battery of tests. More importantly, they lack adequate test sensitivity and specificity, and are susceptible to practice effects which occur when tests are repeated more than once [7].

BrainCheck is an alternative solution to mTBI assessment that is rapid and self-administered. BrainCheck Sport battery is a computerized neurocognitive test that assesses various areas of cognition, such as attention, processing speed, coordination, and executive functioning. Its diagnostic accuracy was previously validated as a testing method for traumatic brain injuries and it is classified as a diagnostic aid by the FDA [8]. We also evaluated the differences between 10 pediatric participants with mTBI diagnosed by physicians at the Emergency Department and 126 control subjects at a local high school on their Braincheck battery assessments and utilized the subsequent metrics to build a logistic regression model that distinguishes mTBI from non-mTBI participants. Receiver operator score (ROC) analysis of our logistic regression model found a sensitivity of 84% and specificity of 81%, which confirmed Braincheck’s potential in assessing cognitive function after brain injury, while also providing a method for tracking recovery remotely from a patient’s home. Thus, the primary objective of this study is to assess the utility of BrainCheck as a diagnostic tool in evaluating mTBI in the pediatric population.

## Materials and methods

This study was approved by the Baylor College of Medicine institutional review board. It took place at the emergency department (ED) of Texas Children’s Hospital (TCH) in Houston, TX and Morton Ranch High School in Katy, TX. We recruited 136 participants in this study, which includes 126 participants in the control group and 10 participants in the mTBI group. Subjects were included in the study if they received an official physician’s diagnosis of mTBI. The recruited volunteers were either enrolled in athletic programs at Morton Ranch or patients at the TCH ED. For the control group we used baseline tests from healthy Morton Ranch athletes, aged 13 to 18 years. One outlier had an extremely high score in the Digit Symbol Substitution Task was removed. After excluding participants who had a history of mTBI or malingered, we were left with 122 participants in the control group and 9 participants in the mTBI group.

### Test Administration

mTBI participants were administered Version 3 of the BrainCheck Sport battery on the iPad within 96 hours of injury by an ED nurse if recruited at TCH or by an athletic trainer if recruited by Morton Ranch. Control participants, composed of Morton Ranch athletes, were administered the BrainCheck test by a research coordinator prior to the athletic season. Prior to the actual BrainCheck test battery, mTBI participants took the Post mTBI Symptom Scale derived from the SCAT3.

### Test Measures

A short description of each assessment that comprises the BrainCheck battery are listed in Table 1. A more detailed description provided of the battery has been described previously [8].

**Table 1.**
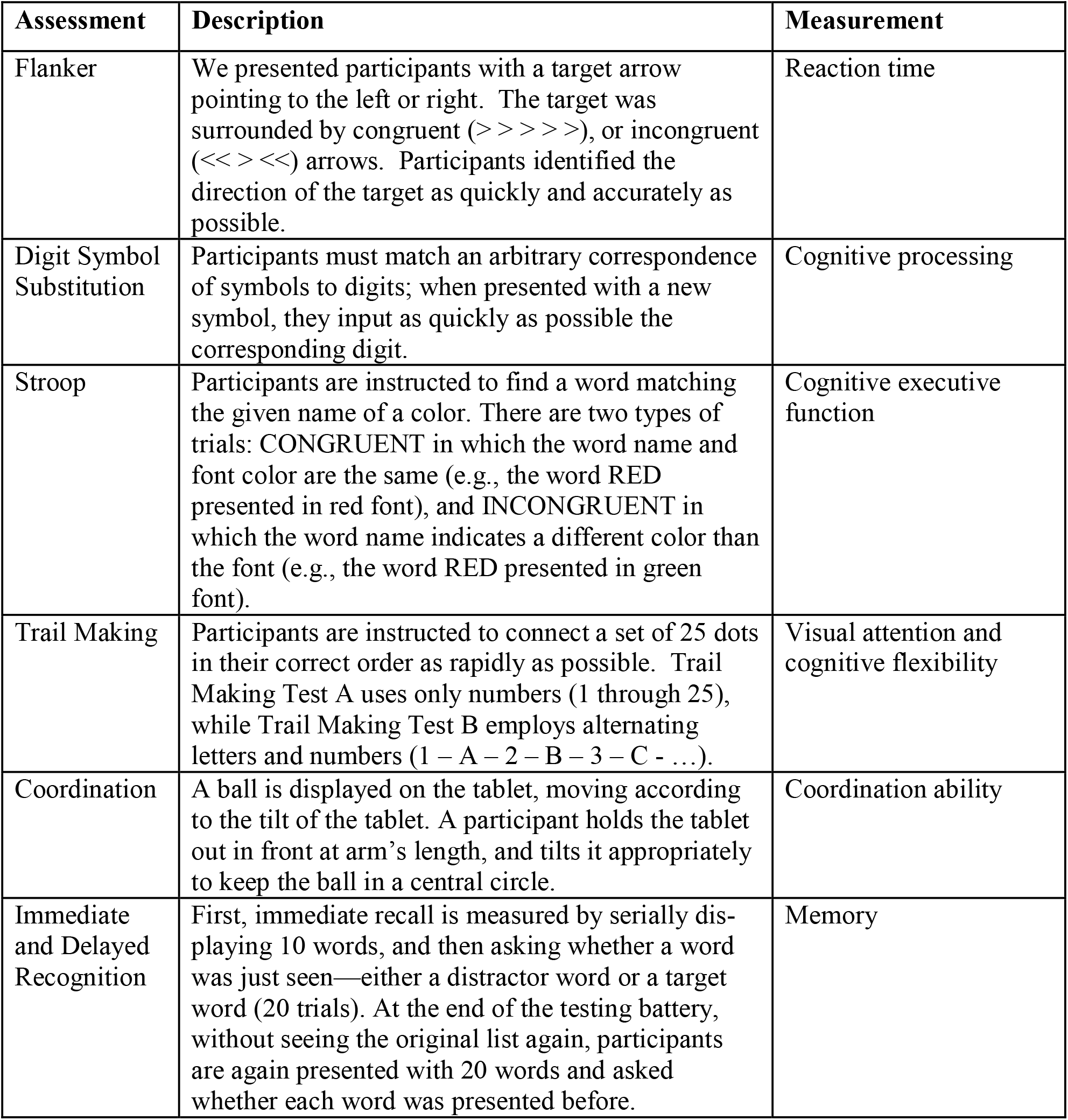
Neuropsychological assessments in BrainCheck battery

### Statistical Analyses

Chi-square tests and Analysis of Variance (ANOVA) were used to examine differences in age and gender in the mTBI and control groups. Independent sample t-tests were used to compare groups on each assessment metric and the Hochberg test was used to adjust p values for multiple comparisons. For each kind of assessment, only the most significant metric was used in our logistic regression model. Sensitivity and specificity of the overall battery was evaluated using receiver operating characteristic (ROC) curve analyses with a cutoff score that represented the maximum values sensitivity and specificity could reach. Data analysis was performed using the R statistical programming language in RStudio version 3.3.1.

## Results

### Demographics

The overall gender distribution was unbalanced because most athletes participated in the study were males (Figure 1). Enrolled participants aged from 8 to 26 years (Figure 2). Nevertheless, there was no statistically significant difference in gender between patients and controls (X-squared (1) = 0.005, p = −0.94). Similarly, no significant difference in age was detected between patients and controls (F(1, 130) = 0.016, p = 0.9).

**Figure 1.**
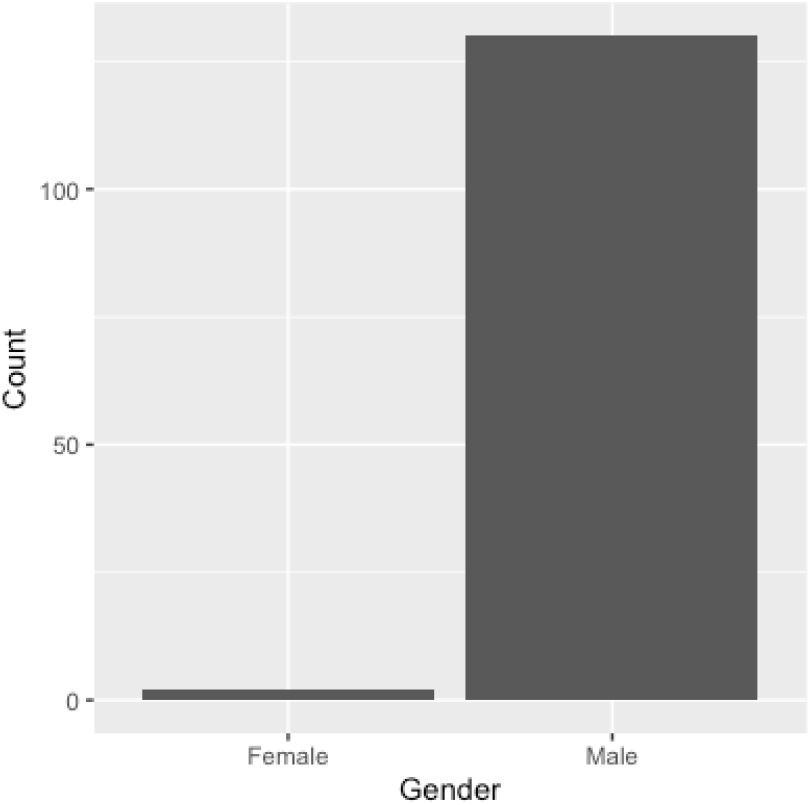
Gender distribution of participants

**Figure 2.**
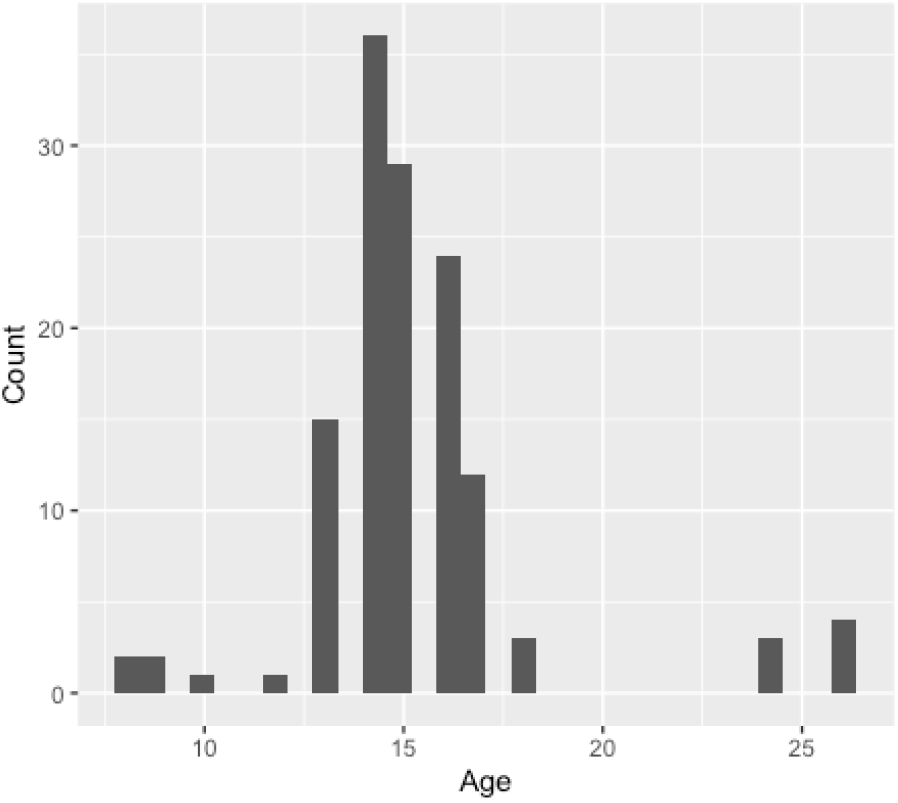
Age distribution of participants

### Assessments scores on mTBI

The Braincheck assessments evaluate different cognitive domains of patients. As shown in Figure 3 and Table 2 below, we found that mTBI and control participants showed significantly difference on raw scores of delayed recall (t_61.5_ = 4.409, p<0.01), immediate recall (t_52.4_ = 4.8192, p<0.01), the Stroop test (t_61.6_ = −4.693, p<0.01), and the Coordination test (t_79.8_ = 3.7075, p<0.01). In contrast, no difference was detected on the Trail Making test (t_72.4_ =-0.64, p=0.52 for trails A, t_66.7_ = −0.83, p = 0.4 for trails B), the Flanker test (t_62.8_ = 2.0934, p = 0.056) and the Digit Symbol Substitution test (t_45_ = −2.0879, p = 0.056).

**Figure 3.**
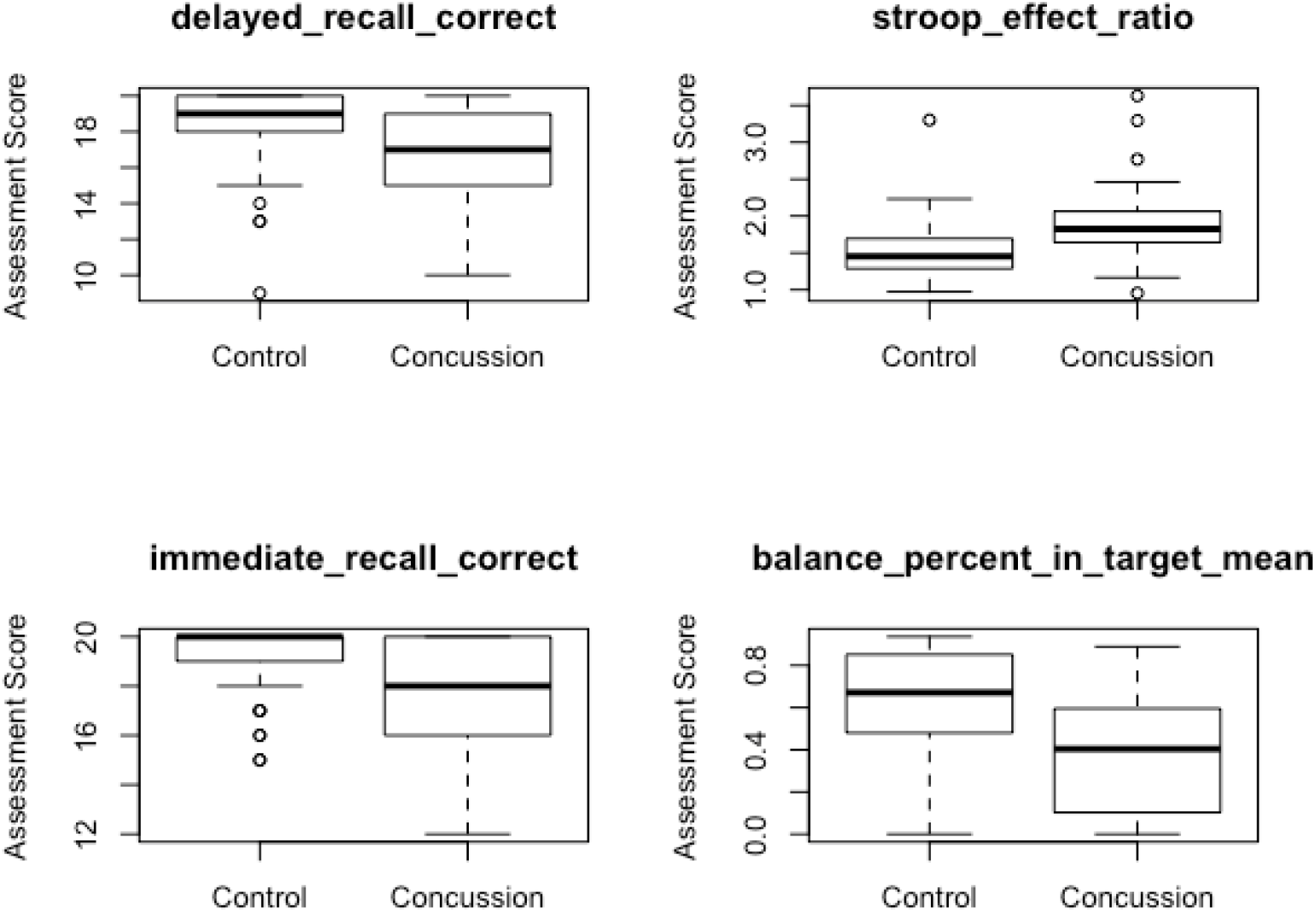
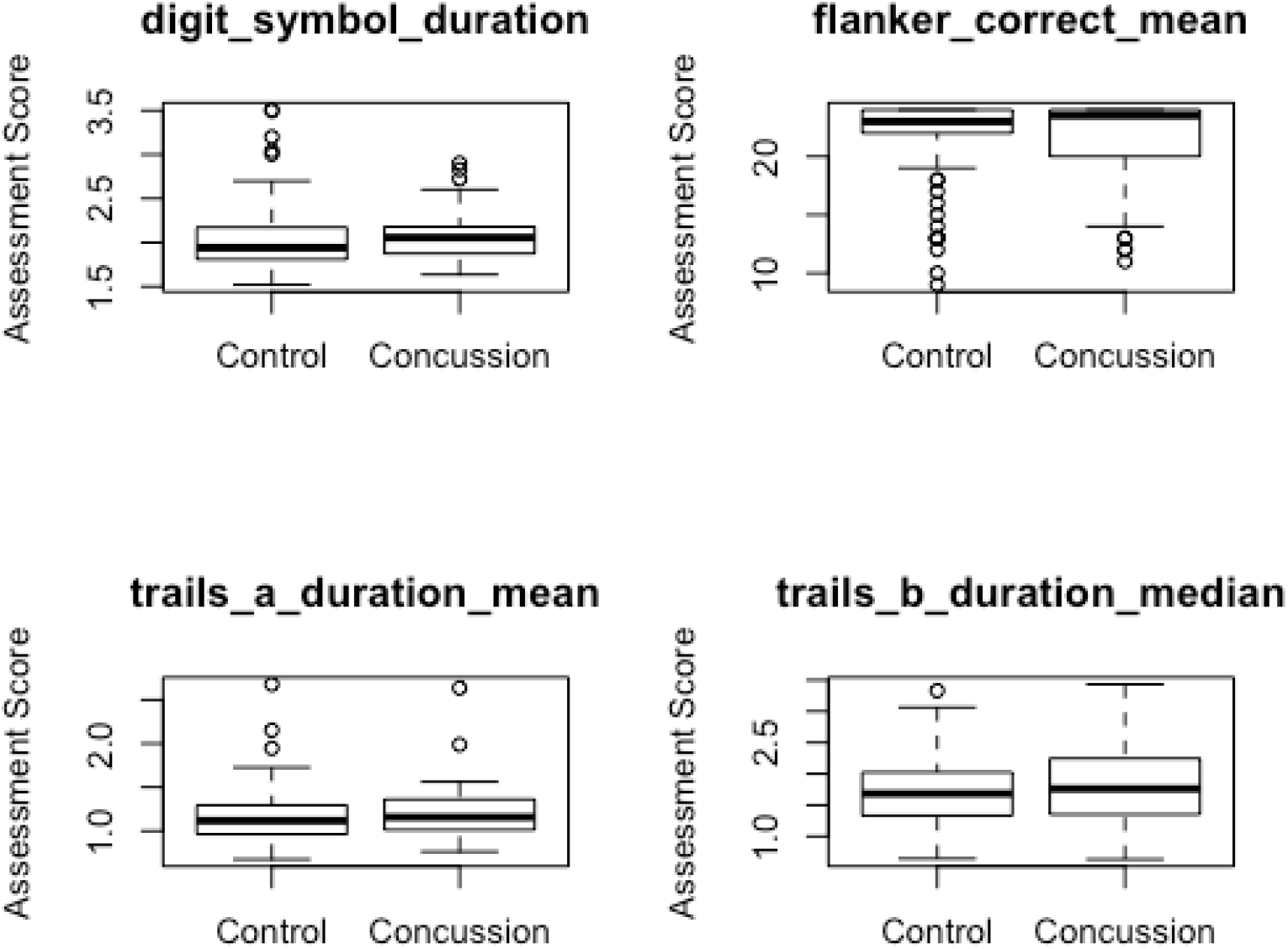
Boxplots of Braincheck assessments metrics for the control group and the mTBI group.

**Table 2.**
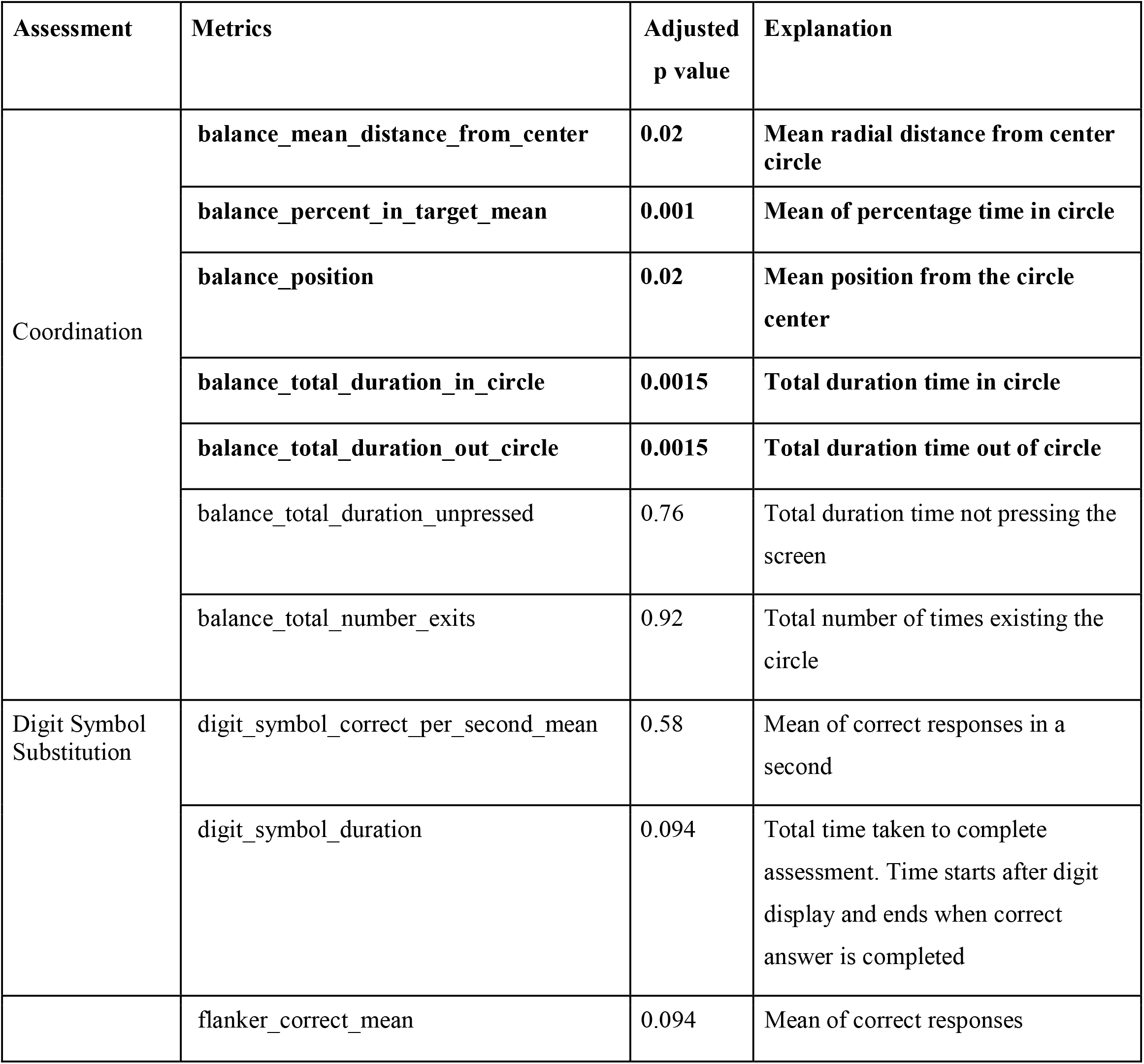
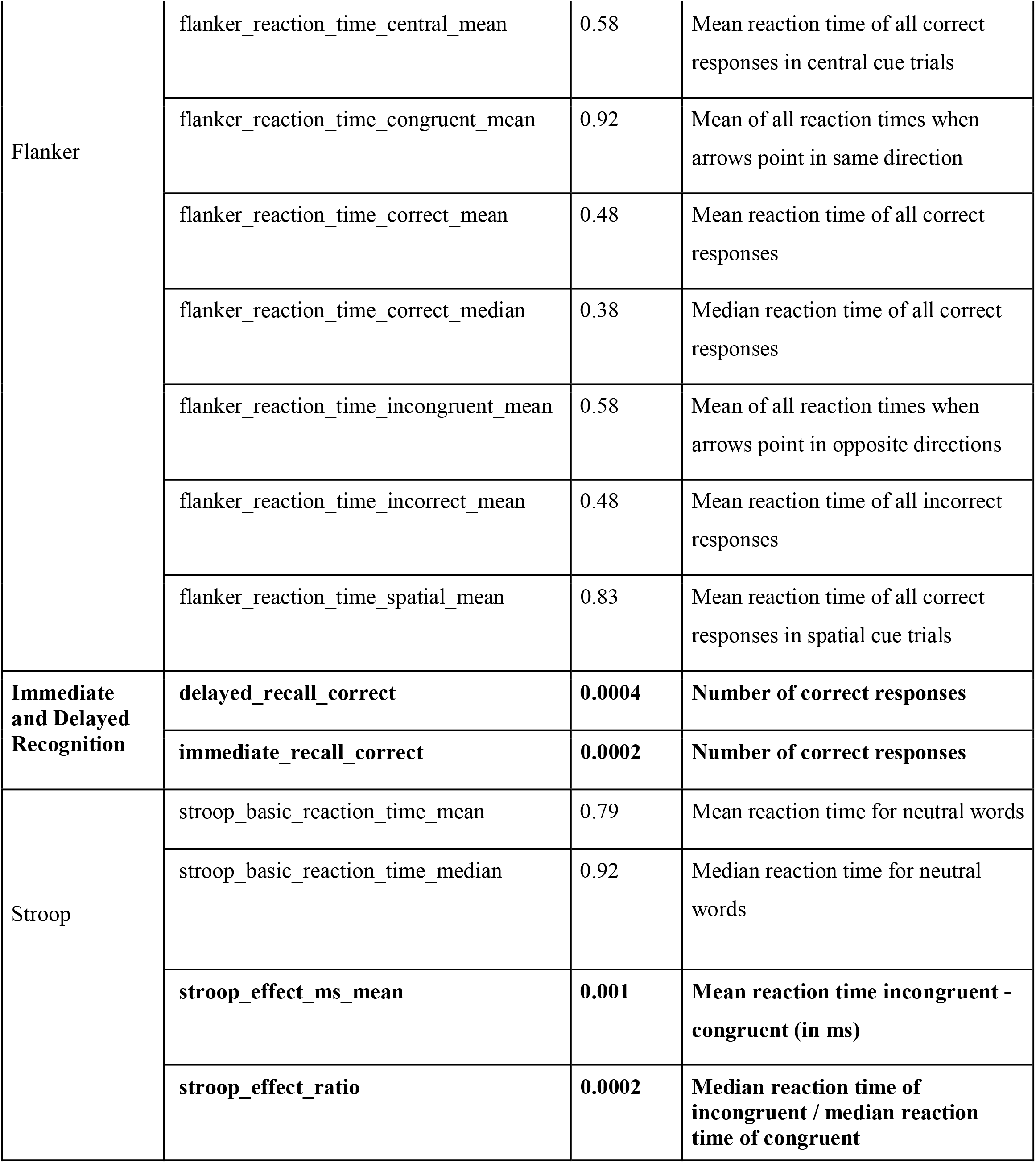
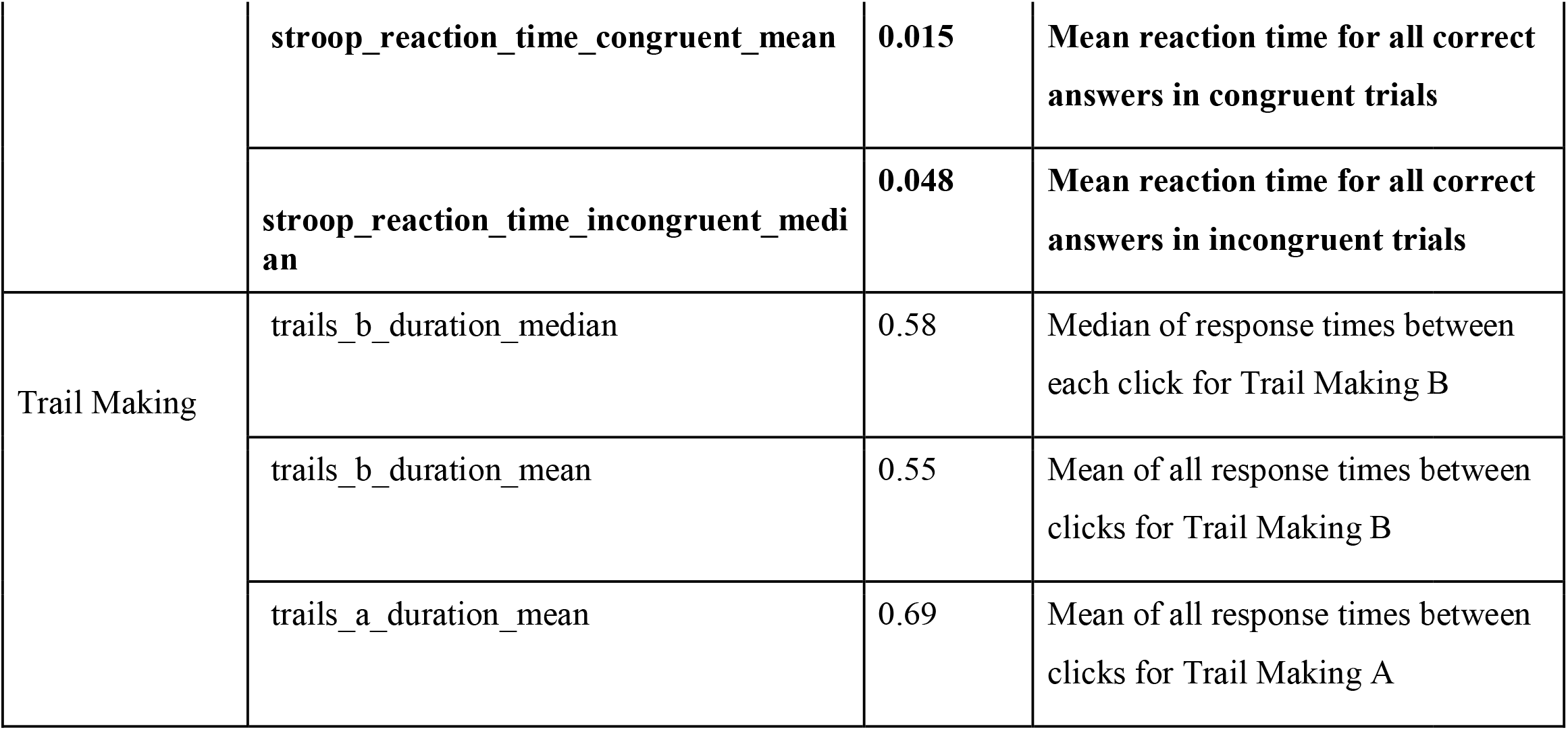
Benjamini-Hochberg adjusted P value for assessment comparison between mTBI group and control group. Statistically significant metrics are listed in bold.

Our results demonstrate that objects that suffered from mTBI exhibited worse performance in Braincheck’s assessments of memory, executive and coordination on the battery, which are consistent with studies conducted golden standard neuropsychological tests [9]-[11]. In addition, they showed no significant deterioration in their cognitive performance and attention, which has also been observed in previous studies [12,13].

### Logistic regression

In our logistic regression model, 30 observations were deleted due to missing data, therefore it utilized 92 of the participants in our control group and 9 of the participants in our mTBI group. We used "delayed_recall_correct", "stroop_effect_ratio", "immediate_recall_correct", "balance_percent_in_target_mean”, "digit_symbol_duration", “flanker_correct_mean”, “trails_a_duration_mean”, "trails_b_duration_median" as factors in our model. Based on the logistic model, the Stroop test, the Immediate Recall test, the Coordination and Trail-making were statistically significant at, while the rest were not statistically significant. Here, we used receiver operating characteristic (ROC) analysis methods to determine the optimal threshold for the logistic regression model so that the model could achieve the maximum sensitivity and specificity to distinguish between control and mTBI group. We found that the BrainCheck Sport battery could achieve a sensitivity of approximately 84% and a specificity of 81% when we chose 0.3 as the optimal decision threshold (Refer to ROC curve in Figure 4). In other words, if the model predicted probability for a subject is greater than 0.3, the subject is categorized to the mTBI group.

**Figure 4.**
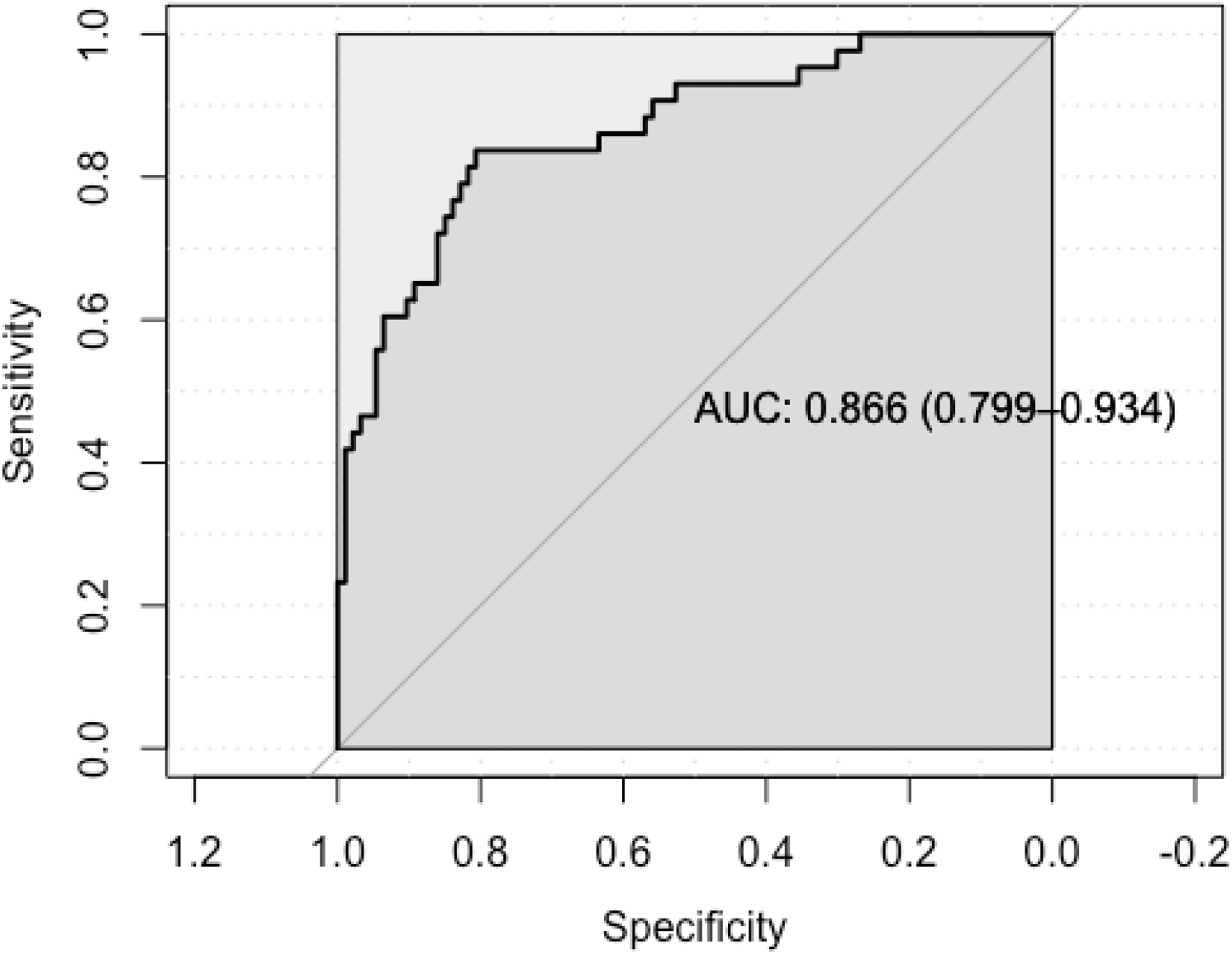
ROC curve of logistic regression.

## Discussion

The findings of this study demonstrated that BrainCheck Sport is able to discriminate mTBI patients from the control group on some battery assessments, and achieves decent levels of sensitivity (84%) and specificity (81%) when classifying mTBI. This is achieved by evaluating the cognitive function, memory, and coordination of the participants under tests. The results reflect that Braincheck Sport would be as effective of a diagnostic aid in the pediatric population as it would be in the adult population [8].

Our results demonstrate that after mTBI, patients experience deficits on immediate and delayed recall tests; balance and coordination tests; and the Stroop color tests which represent deficits in cognitive domains consisting of short term memory; coordination; and executive function, specifically inhibition and attention, respectively. These are typical symptoms observed after concussions. We did not observe degradation in patient performance on trail making; digit-symbol substitution; or the Flanker tests which represent cognitive domains consisting of executive function, specifically, cognitive flexibility; information processing and speed; and visual attention, respectively. As suggested by previous studies, not all cognitive domains of concussion patients will be affected [12,13]. However, De Beaumont et al. [14] found that athletes with concussion history performed worse in the Flanker Test. This might be due to the repeated injuries these athletes sustained and became significant over time. In our study we excluded participants with mTBI history, and participants were relatively young. Therefore, it is likely their symptoms differ from those who have experienced repeated concussions over time and are older. On the other hand, the majority of our participants are only assessed within 48 hours after injury leaving time for symptoms to possibly resolve. Further studies should control for time of administration after injury more stringently by restricting the timeframe to within 24 hours or less.

It is important to keep in mind that our participants were from an atypical population. The control group used was composed of athletes and was not fully representative of the pediatric population. Since the mTBI experimental group included both sports related and non-sports related mTBIs, a control group with similar demographics would have contributed to a more balanced design. We also sacrificed statistical power with a smaller mTBI group, which may have resulted in less separation of groups in our significance tests. A larger experimental group with more individuals in the mTBI group would likely give more robust results. It is possible to have a large range of symptoms after concussion with a range of cognitive impairment which is likely present in our data. Having a larger dataset would minimize the effects of this variation within the concussion population. In addition, both our experimental and control groups were composed of only a few females. Such gender differences in mTBI injury severity have been reported in previous literature [15]. To investigate whether there are gender differences in the neurocognitive measures examined in the BrainCheck battery, future research should utilize a more evenly distributed ratio of males to females in the participants pool. Additionally, despite being a statistically significant predictor, the coordination test occasionally suffered from software malfunctioning during testing, and resulted in several cases of missing data in both the experimental and control groups. As a retrospective study, it is hard to determine if all cases of malfunctioning were properly documented and fully taken into consideration.

Currently there are a variety of different test batteries and screeners used in clinical practice to aid in the diagnosis of mTBI. BrainCheck provides another option among the computerized neurocognitive tests by providing a shorter, gamified test battery that attempts to comprehensively examine and assess cognitive functioning after brain injury. Here we demonstrate BrainCheck performs well in detecting and classifying mTBI patients, which may be useful as a diagnostic aid.

## Data Availability

Data may be made available by contacting the corresponding author and with a data use agreement.

## Abbreviations

mTBI: :mild traumatic brain injury
CT: :computed tomography
ED: :emergency department
ANOVA: :Analysis of Variance
ROC: :Receiver operator score
TCH: :Texas Children’s Hospital

## Acknowledgments

Funding was provided by BrainCheck, Inc.

## Ethical approval

This study protocol was reviewed and approved by Solutions IRB.

## Consent to participate

The informed consent to participate in the study was obtained from all participants.

## Consent to publication

Not applicable.

## Data Availability

Data may be made available by contacting the corresponding author and with a data use agreement.

## Author contributions

DME and YK contributed conception and design of the study; Data acquisition was performed by BF; SY and BH performed the statistical analysis, under the supervision of RHG. SY, BK and HP wrote the first draft of manuscript, which was revised and approved by BH and RHG.

## Copyright

© The Author(s) 2020.

